# Assessing the impact of periodontal therapy on tooth loss: a register-based longitudinal study in Denmark

**DOI:** 10.1101/2024.06.06.24308349

**Authors:** Eero Raittio, Vibeke Baelum

## Abstract

**Background:** While regular periodontal care is advocated to reduce tooth loss considerably among periodontitis patients, evidence from observational studies is often limited to small single-center studies. This study aims to quantify the effect of periodontal care on tooth extractions using comprehensive Danish register data.

**Methods:** A nation-wide register-based cohort study was conducted, encompassing 40-year-old individuals with incident periodontitis in 2001, tracked through Danish registers until the end of 2021. Receiving any periodontal care was determined annually, and the number of non-surgical tooth extractions serving as the annually varying outcome. G-estimation of structural nested mean models adjusted for time-varying confounding and loss to follow-up were employed to estimate the average treatment effect of periodontal therapy on subsequent tooth extractions.

**Results:** The study included 1,251 40-year-olds with incident periodontitis in 2001. The average follow-up from 2002 onwards was 19.1 years and amounted to 23,878 person-years. On average, participants received periodontal care in 12.1 years (SD 6.3) and lost an average of 1.4 teeth (SD 3.0). G-estimation showed that receiving periodontal therapy in a given year reduced the number of teeth extracted in the following year by 0.03 (95% CI: -0.00; 0.05). The cumulative effect of receiving periodontal therapy for five consecutive years was associated with an average of 0.06 (95% CI: 0.03; 0.11) less extracted teeth, while on average 0.36 teeth were lost in a five-year period (0.072 per year). This protective effect was observed regardless of the baseline severity of periodontitis.

**Conclusions:** Periodontal therapy resulted in a modest reduction in tooth extractions. The effectiveness of periodontal therapy against tooth loss seems to be considerably smaller than indicated by earlier clinical studies.

## INTRODUCTION

The consensus within the dental community underscores the importance of regular periodontal care in reducing adverse disease outcomes, such as tooth loss, in patients with periodontitis.^1–6^ Much of the evidence supporting this consensus comes from observational studies that, while valuable, are often limited by their scale,^2,3,7^ focusing on a single university, hospital, specialist practice or even individual practitioners.^8–10^ These studies consistently report higher rates of tooth loss among patients who do not adhere to recommended periodontal treatment plans, underscoring the potential benefits of regular care.^2,3,7^

Indeed, observational research plays a crucial role in our understanding of the effectiveness of regular periodontal care, primarily because it circumvents the ethical, practical, and financial hurdles of large and long-term randomized controlled trials. Despite their inherent limitations, observational studies can track large populations over extended periods, providing valuable data on long-term outcomes of long-term treatments of relatively slowly progressing chronic diseases like periodontitis. Causal inference from observational data requires careful design and sometimes sophisticated statistical methods.^11,12^ This is particularly true for long-term and time-varying treatments,^13,14^ such as periodontal care, which are influenced by a broad set of time-varying factors including previous treatments, socioeconomic conditions, and the severity and progression of the disease.^2,3,7^ However, such methods have not been applied to investigate the effectiveness of regular periodontal care on reducing adverse disease outcomes, such as tooth loss.^2,3,7^

The aim of the current study was to estimate the effect of receiving periodontal care on the number of subsequent extractions in periodontitis patients over a long follow-up using Danish register data and robust causal inference methods.

## MATERIAL AND METHODS

The data used for the present analyses originate from a large, register-based prospective dynamic cohort study based on Danish register data. All permanent residents have a unique civil personal registration number, which, via the Danish Civil Registration System, allows follow-up and linkage of individual-level information from multiple registers. We generated the cohort by linking information from the Civil Registration System, the Educational Register, the Income Statistics Register, the National Health Insurance Service Register and the Register for Selected Chronic Diseases. People were eligible for entry into the cohort when they reached the age of 20 years in the period from 1 January 1990 to 31 December 2021, were alive at the time of entry, and were permanent residents of Denmark. The study was approved by the Danish Data Protection Agency.

### Sample selection and follow-up

Since 2000, private dental practitioners have had to report the number of teeth present, the number of filled teeth, and the number of decayed teeth for their patients turning 25, 40 or 65 years during the ongoing calendar year^15^. Data are reported to the National Board of Health^15^ where the data have become integrated with the National Health Insurance Service Register. From the National Health Insurance Service Register, we also obtained information on the dental treatments covered by the National Health Insurance scheme for subsidized dental care for all residents in Denmark between 1990 and 2021. This registry thus contains information on all patients, providers, and the services provided insofar as these are publicly subsidized, and regardless of whether a person has additional private insurance or not. Because the register does not contain diagnosis codes, we defined periodontitis based on the periodontal treatment provided, as done in other studies.^16,17^ A person was considered a periodontitis case from the year onwards when the person received their first course of periodontal treatment that involved subgingival/root surface instrumentation in pathological/deepened pockets.

These treatments were identified with treatment codes 1420 (general periodontal treatment), 1425, 1430 (extended periodontal treatment), 1452 (control after general periodontal treatment), 1453 (control after extended periodontal treatment), 1431 (root debridement), 1440 (periodontal surgery), and 1454 (control after periodontal surgery). In the following, we will use the moniker “incident periodontitis” to describe this first occurrence of a course of periodontal treatment that involved subgingival/root surface instrumentation in pathological/deepened pockets. People selected this way might have been exposed to supragingival scaling and/or individual preventive services prior to becoming cases of incident periodontitis, but they had no prior periodontal treatments involving subgingival instrumentation recorded.

For the purpose of this study, we selected people fulfilling all the following: 1) turned 40 years in 2001, 2) had incident periodontitis in 2001, 3) had a dental examination which included the reporting of the number of teeth present, the number of filled teeth, and the number of decayed teeth for the year 2001, and 4) had lived permanently in Denmark between 1997 and 2002. All individuals thus selected were followed from 1997 until the earliest of the following events: the end of 2021, the year of death, or permanent relocation abroad (Figure 1).

**Figure 1.**
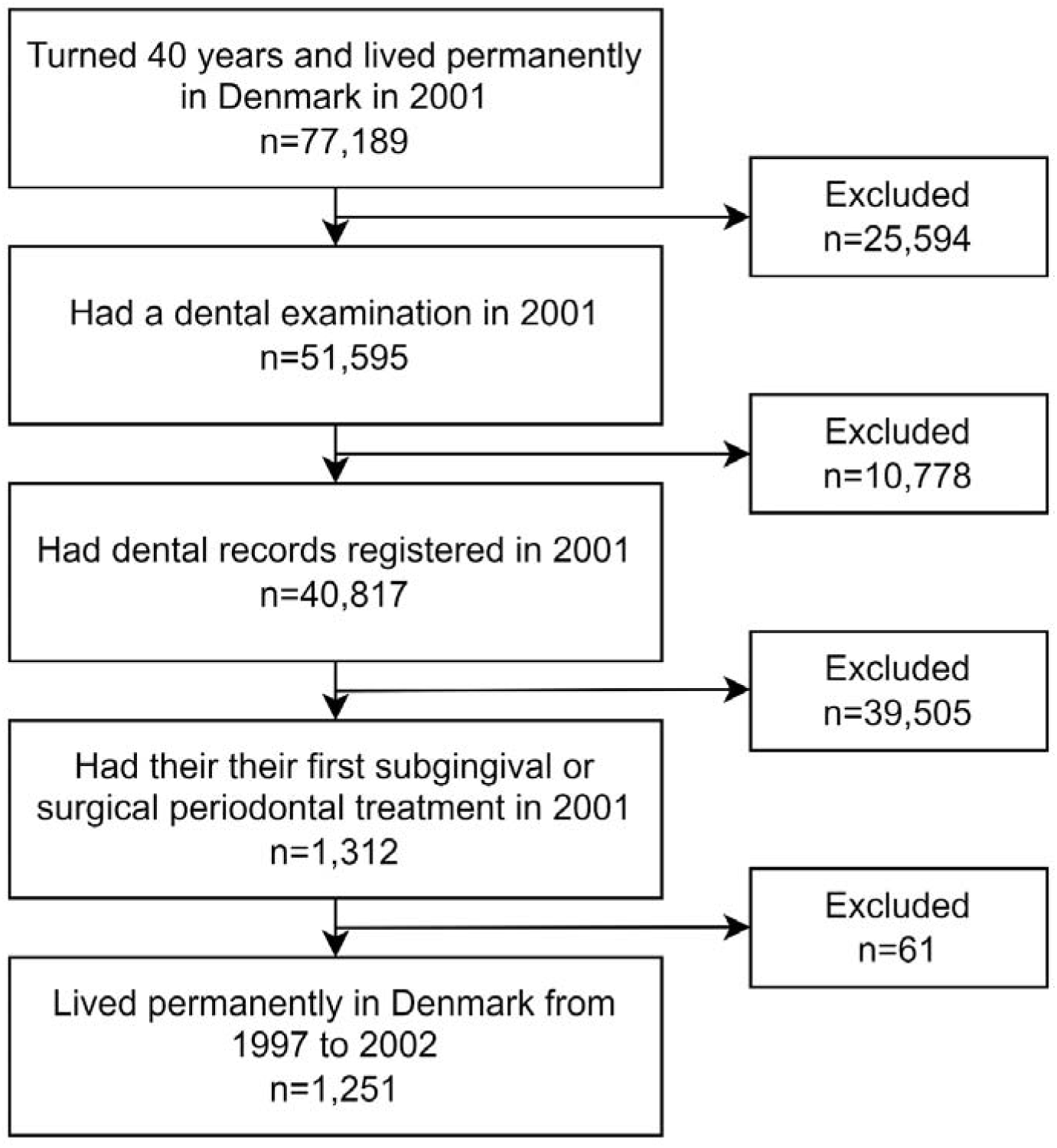
Flow chart of sample selection process.

The rationale behind these criteria was to achieve a sample of 40-year-old individuals with incident periodontitis in 2001 who could be followed up for tooth extractions after having been observed to be periodontitis free (no recording of subgingival or surgical periodontal treatments) for a period extending at least back to 1997, and possibly back to 1990, while providing information about the numbers of teeth, filled teeth, and decayed teeth from 2001.

### Exposure and outcome

Any person selected as outlined above was defined to be exposed in the calendar year if the person had received (yes/no) periodontal care in the form of individualized prevention, or any supragingival, subgingival, or surgical periodontal treatment according to the National Health Services Register. The number of non-surgical tooth extractions in each calendar year was determined based on the treatment codes for tooth extractions and used as an outcome.

### Covariates

The time-invariant covariates considered in the analysis (Figure 2, Supplementary material) were the 2001 recordings of gender, origin (immigrants, descendants of immigrants, Danish origin), number of teeth present (0-28), number of decayed teeth (0-28), number of filled teeth (0-28), and presence (yes/no) of severe periodontitis, defined by the receipt of root debridement or periodontal surgery (treatment codes 1431, 1440).

**Figure 2.**
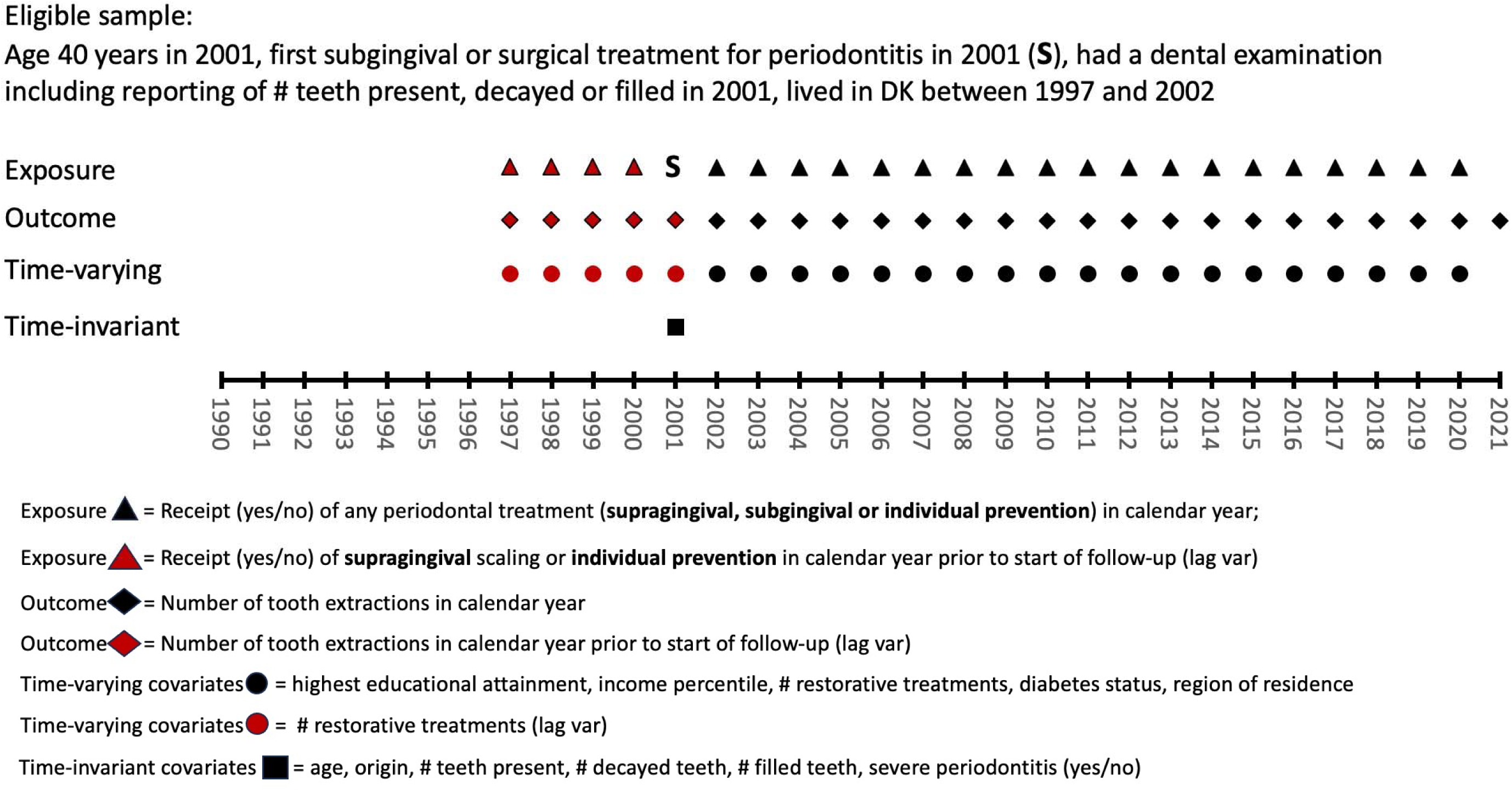
Overview of the data structure underpinning the analyses.

Annually time-varying covariates (Figure 2, Supplementary material) considered were the region of residence (North Jutland, Central Jutland, Southern Denmark, Zealand, Greater Copenhagen), the highest educational attainment (eight levels), the income percentile, the number of restorative treatments in a calendar year, and type 1 or type 2 diabetes status, which turned from 0 to 1 in the year the patient got diabetes according to the Danish National Diabetes Register (Supplementary material).

### Statistical analysis

G-estimation of structural nested mean models^13,14,18–21^ was used to estimate the average treatment effect of having received periodontal therapy in a calendar year on the number of extracted teeth in the subsequent years. Traditional statistical analysis methods are not suitable for such causal inference due to the presence of time-varying exposure and outcome, and time-varying confounding occurring when confounding is affected by earlier exposure or outcome status, or both. This can be shown with a directed acyclic graph (DAG) (Figure 3), which illustrates a simplified version of our analytical design involving only three time points, labeled 1-3. Suppose we have a time-varying binary exposure E (receiving periodontal therapy), and an outcome O (the number of tooth extractions), measured at each of the years t=1,…., T. The exposure and outcome in a given year will be influenced by 1) previous exposure and outcome status, 2) time-invariant confounders C (e.g., gender or baseline disease severity), 3) time-varying confounders W (e.g., income or education) affecting exposure and outcome with or without lag, 4) time-varying confounders V, affecting exposure and outcome only with lag, such as treatments given at the same time as the exposure or the outcome. Note also that time-varying confounders are affected by previous exposure and outcome status, and that from the last time point, only outcome data is used.

**Figure 3.**
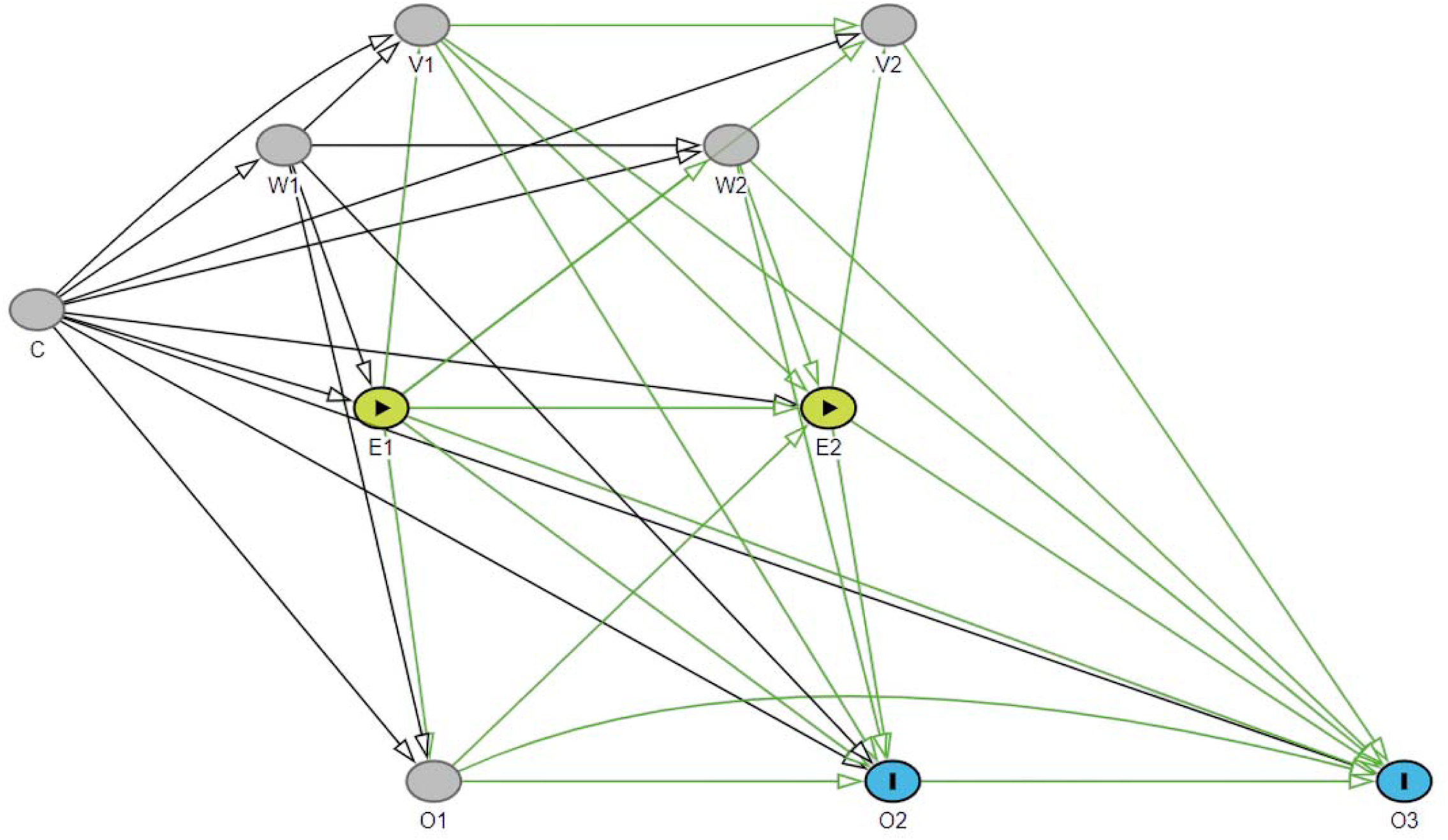
A directed acyclic graph showing a simplified version of analytical design. E is the exposure of interest; O is the outcome; C represents time-fixed confounders; W and V represent time-varying confounders affecting with and without lag (W) or only with lag (V). Numbers (1, 2 or 3) refer to time point. Green paths are causal paths from exposure to outcomes of interest.

We are interested in measuring the effect of E1 on O2, E1 on O3, and E2 on O3. The DAG presented in Figure 3 indicates that to estimate the effect from E1 to O3, one should and should not adjust for time-varying confounders (W2, V1, V2) and previous outcomes (Y2, Y1) because they are both mediators and confounders on the causal pathways from E1 to O3 (green). In line with the principles of causal inference, if one adjusts for a mediator, it induces a collider bias and if one does not adjust for confounding, there will be bias from uncontrolled confounding. In such cases the use of traditional statistical methods leads to biased results.^13,14,19^ The supplementary material explains how the g-estimation of structural nested mean models handles the situation by sequentially estimating the counterfactual outcomes when the exposure is hypothetically manipulated.^13,14,18–21^

We therefore used the g-estimation of structural nested mean models by applying variable specifications as outlined in the following. In line with the simplified DAG (Figure 3), our exposure was the time-varying binary recording of receiving periodontal therapy (yes/no) in a calendar year from 2002 (E1) to 2020 (E19), and our outcome was the number of extractions in a calendar year from 2003 (O2) to 2021 (O20). Gender, origin, baseline number of teeth, baseline number of decayed teeth, baseline number of fillings, baseline disease severity were the time-invariant confounders (C) measured in 2001 (T=0); while educational level, income, municipality/region and diabetes status were considered time-varying confounders with and without a 1-year lag (W); while the number of restorations was considered a time-varying confounder affecting only with a 1-to-5-year lag (V). Additionally, previous outcome and previous exposure status were allowed to affect subsequent outcome and exposure status with a 1-to-5-year lag already from the beginning of the follow-up (2002, T=1) (Supplementary material). In the analyses, continuous covariates with sufficient variability – in our case income and number of fillings - were allowed to have non-linear relationships with the outcome and the exposure using natural cubic splines. Additionally, censoring weights (death/move abroad) were generated using the same adjustment sets.

Utilizing the above-mentioned model specifications and the gesttools^20^ package in R (Supplementary material), we conducted g-estimation of structural nested mean models to estimate the time-varying average treatment effect of receiving periodontal therapy at any exposure time on the number of extracted teeth in the ensuing one to five year period. Three different outcome specifications were used. First, we treated the number of extractions within a calendar year as a continuous variable and employed linear regression in the g-estimation. Even though the linear modeling of the number of extracted teeth in a calendar year most likely violates the assumptions underpinning the linear regression, it allows us to estimate what would be the effect of receiving periodontal therapy each year over a 5-year-period compared to not receiving periodontal therapy in any year over the 5 years.^20,21^ We also ran analyses considering the outcome as a count variable, or as a dichotomized variable, i.e., having at least one tooth extraction within a calendar year. In both cases, gamma regression with a log link function was used instead of linear regression.^18,20^ In these cases the effect modelled is either the ratio of the expected numbers of tooth extractions within a calendar year in one to five years ahead or the ratio of the probability of having at least one tooth extraction within a calendar year. Again, we estimate the time-varying average treatment effect of receiving periodontal therapy at any exposure time on the number of extracted teeth in the following five years.

Considering that the effect might differ based on the baseline severity of the disease, we also allowed baseline disease severity to modify the effect of receiving periodontal therapy. 95% confidence intervals (CIs) for the estimates were generated by bootstrapping the dataset 500 times. In addition to basic descriptive statistics, we report the findings from the propensity score model used, indicating how the included variables were associated with receiving periodontal therapy within a calendar year.

### Sensitivity analyses

As we did not determine the actual temporal relationships between the dental treatments received within a given year, the DAG in Figure 3 represents just one possible data generation process. A sensitivity analysis was therefore conducted based on an alternative DAG, in which the arrows from E to O and from E to V were reversed in each year (Figure 4). This means that instead of assuming that other treatments (V) and extractions (O) in same year are caused by periodontal care (E), they are considered determinants of receiving periodontal care in the same year. Therefore, they should be included in the models estimating the effects originating from the same year, and not omitted as we did in the primary analyses. These analyses served to indicate the sensitivity of the results to the specification of the unknown temporal relationships among recordings from the same year.

**Figure 4.**
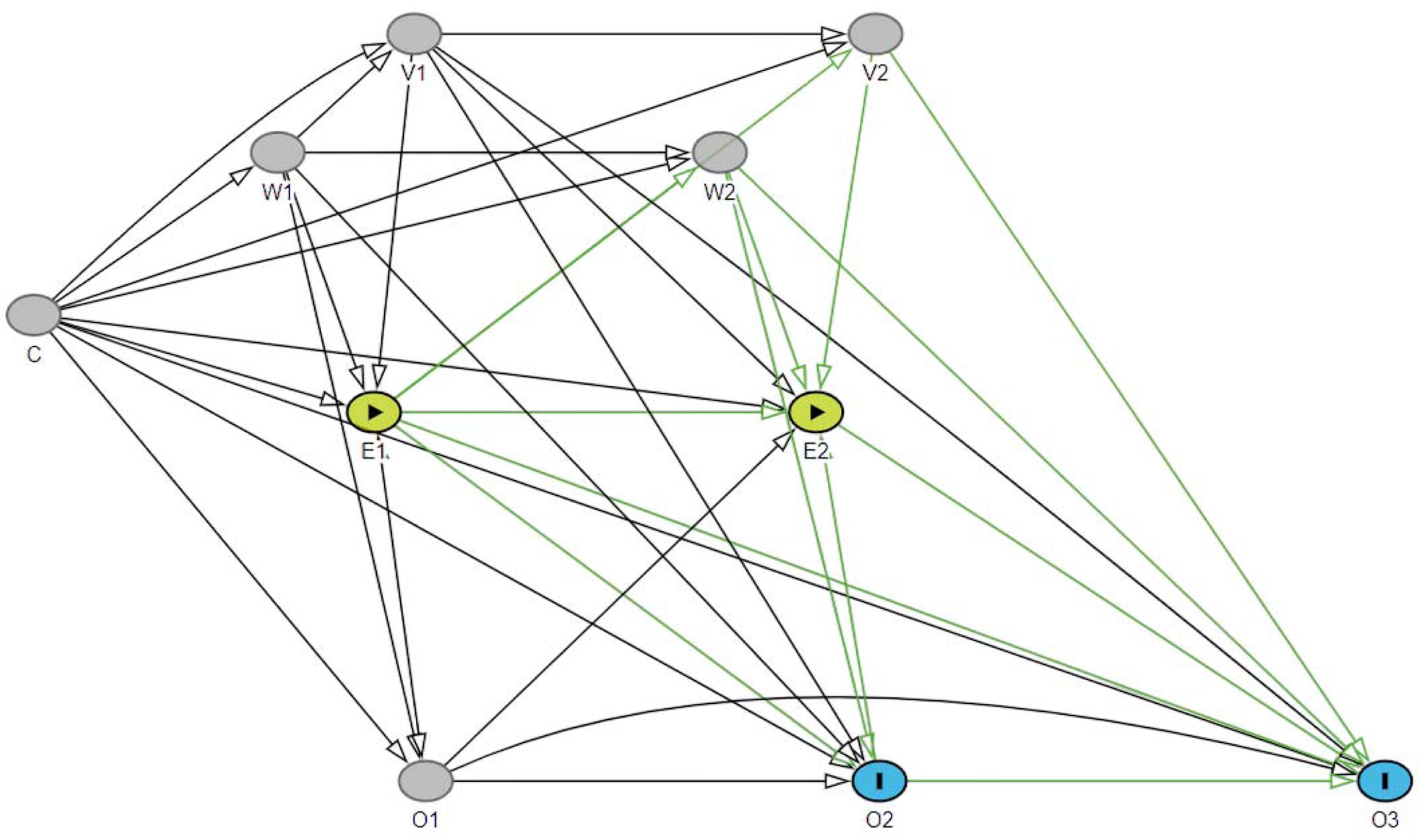
An alternative DAG. E is the exposure of interest; O is the outcome; C represents time-fixed confounders; W and V represent time-varying confounders affecting with and without lag (W) or only with lag (V). Numbers (1, 2 or 3) refer to time point. Green paths are causal paths from exposure to outcomes of interest.

## RESULTS

The final sample of 40-year-old individuals with incident periodontitis in 2001 included 1,251 persons (Figure 1), who were followed for 30,133 person-years from 1997 to 2021, or up to censoring due to death or moving abroad after 2002. The average duration of the follow-up from 2002 onwards was 19.1 years and amounted to 23,878 person-years.

A little over half of the sample were men and about one half had severe periodontitis at baseline (Table 1). The vast majority were of Danish origin and had a complete dentition (28 teeth) at baseline. At baseline (2001) the median number of filled and decayed teeth was 12 (IQR=8-16) and 0 (IQR=0-2), respectively. The sample had a higher average income than the general population (median income percentile over 50), and over two-thirds of the sample had vocational education as their highest educational level.

**Table 1.** Characteristics of the analyzed sample in 2002 and 2020.

|  | **2002** | **2020** |
| --- | --- | --- |
| **Baseline characteristics** | **n = 1 251** | **n = 1 140** |
| Men, n (%) | 686 (55) | 610 (54) |
| Severe periodontitis, n (%) | 646 (52) | 592 (52) |
| No. of teeth, median (IQR) | 28 (28, 28) | 28 (28, 28) |
| No. of filled teeth, median (IQR) | 12 (8, 16) | 12 (8, 16) |
| No. of decayed teeth, median (IQR) | 0 (0, 2) | 0 (0, 2) |
| Danish origin, n (%) | 1 149 (92) | 1 047 (92) |
| **Time-varying characteristics** |  |  |
| No. of restorations in a calendar year, median (IQR) | 0 (0, 1) | 0 (0, 1) |
| Income percentile, median (IQR) | 69 (50, 84) | 68 (43, 82) |
| Type 1 or 2 diabetes, n (%) | 10 (0.8) | 102 (8.9) |
| Educational level, n (%) |  |  |
| Primary school | 352 (28) | 263 (23) |
| Vocational | 508 (41) | 481 (42) |
| Short-cycle higher education | 70 (5.6) | 69 (6.1) |
| Medium-cycle higher education | 152 (12) | 165 (14) |
| Other (4 levels) | 169 (13) | 162 (14) |
| Region, n (%) |  |  |
| Northern Jutland | 106 (8.5) | 108 (9.5) |
| Central Jutland | 288 (23) | 261 (23) |
| Southern Denmark | 256 (20) | 242 (21) |
| Capital region | 408 (33) | 342 (30) |
| Zealand | 193 (15) | 187 (16) |

Figure 4 shows that 60-75% of the individuals received periodontal care in the calendar year from 2002 to 2021. The percentage slightly decreased over the years. On average, a person had 12.1 (SD 6.3) years in which they received periodontal care. The average number of extractions in a calendar year increased from approximately 0.05 to 0.10 during the follow-up (Figure 5), being 0.07 on average. Over follow-up, 1,715 teeth were lost, amounting to an average of 1.4 (SD 3.0) teeth lost per person.

**Figure 5.**
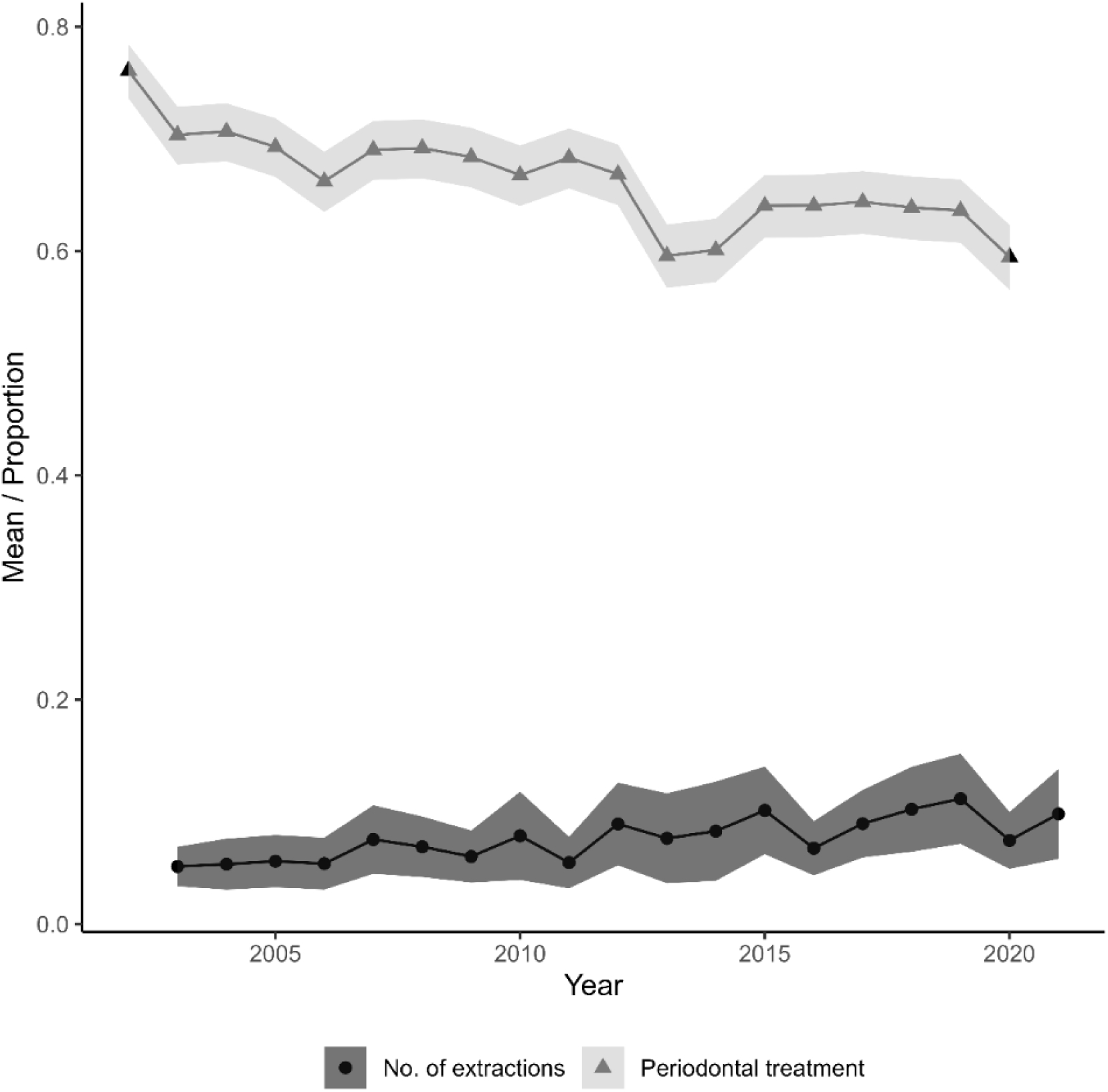
The average number of extractions within a calendar year from 2003 to 2021 (outcome) and the proportions of individuals who received periodontal treatment within a calendar year from 2002 to 2020 (exposure).

Receiving periodontal therapy within a calendar year was strongly associated with having received periodontal therapy in the previous five years, e.g., having received periodontal therapy in the previous year was associated with 5.52 higher odds of receiving periodontal therapy next year (Table S1). Having received extractions in the previous five years was associated with a slightly lower probability of receiving periodontal therapy in the calendar year. Other variables associated with receiving periodontal therapy within a calendar year were income splines, region, education, gender, origin, disease severity at baseline and calendar year.

G-estimation showed that receiving periodontal therapy in a calendar year resulted in the number of extracted teeth in the following year being lower (effect = - 0.03 teeth, 95% CI: - 0.05; 0.00, Figure 6A) than seen among those not receiving periodontal therapy. The effect on the number of extracted teeth two years later was slightly smaller than the effect on the number of extracted teeth three to four years after treatment, and the effect of receiving periodontal therapy in a calendar year on the number of teeth extracted five years later was close to null (effect = 0.02, 95% CI: -0.01; 0.05, Figure 6A). Because these effect estimates originated from the linear models, we can estimate that receiving periodontal therapy in five years a row compared to not receiving any periodontal therapy is associated with an average of 0.06 (95% CI: 0.03; 0.11) less extracted teeth. The effect was practically similar whether people had severe or non-severe periodontitis at baseline (Figure 6B).

**Figure 6.**
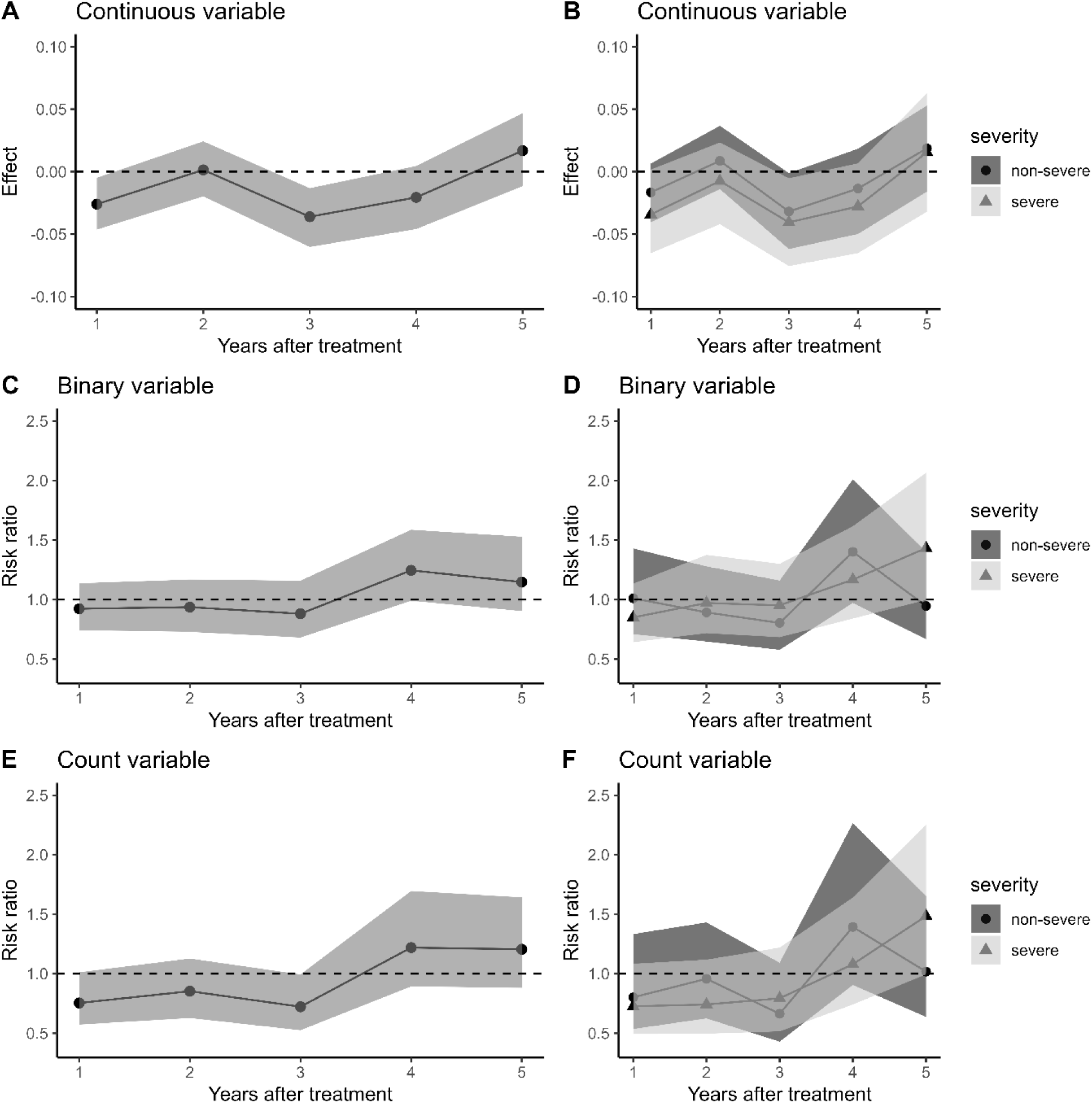
The effect (difference between non-treated and treated) of receiving periodontal treatment within a calendar year on the number of extracted teeth (A, B, E, F) or probability of at least one tooth extraction (C, D) one to five years later. The right panel shows the effect of receiving periodontal treatment within a calendar year in those with severe or non-severe condition at the baseline.

On the risk ratio scale, the effect of periodontal therapy amounted to a risk ratio of 0.92 (95% CI: 0.74; 1.14) indicating a lower risk of receiving a tooth extraction (Figure 6C) and a 0.75 (95% CI: 0.57; 1.01) lower ratio of the expected number of tooth extractions in the year following periodontal therapy (Figure 6E) among the treated. The small protective effect of receiving periodontal therapy within a calendar year lasted three years (Figures 6C,E), and the effect was close to null or even associated with higher risk of tooth extractions four to five years later. Again, the effect was practically similar whether people had severe or non-severe periodontitis at baseline (Figure 6D,F). Results were practically identical when variables were operationalized in line with the alternative DAG (Figure S1).

## DISCUSSION

The findings of this register-based study indicate that the effect of receiving periodontal therapy in a calendar year reduces the number of extracted teeth by 0.03 (95% CI: -0.00; 0.05) teeth in the following year. This corresponds to a ratio of the expected number of tooth extractions in the year following periodontal therapy of 0.75 (95% CI: 0.57; 1.01) indicating that if the untreated would expect one tooth extraction the corresponding figure would be 0.75 for the treated. Similarly, the risk ratio for a tooth extraction in the year following periodontal therapy was 0.92 (95% CI: 0.74; 1.14), indicating an 8% lower risk among the treated. The protective effect of periodontal treatment seemed to disappear after three years, and we can estimate that receiving periodontal therapy for five years in a row compared to not receiving any periodontal therapy was associated with an average of 0.06 (95% CI: 0.03; 0.11) less extracted teeth. The effect of periodontal therapy was practically similar whether people had severe or non-severe periodontitis at baseline.

Even though we were able to use robust causal inference methods and long-term, rich, and nationally representative Danish data, this study is not without limitations. Because treatment and dental health records in the National Health Insurance Service Register are mainly gathered for administrative purposes, there may be variation in how the number of teeth, number of filled teeth, and number of decayed teeth are recorded, or in how the legal guidance and definitions of indications for the treatment codes have been applied. However, because the potential misclassification or measurement error are not likely to depend strongly on our exposure or outcome, the implications are presumably relatively small for the results of the analyses. As with all observational research, particularly register-related research, there is always some degree of unmeasured confounding. Factors related to tooth loss and adherence to periodontal therapy, which we were not able to consider, may have biased some of our effect estimates towards null and some away from null. For instance, smoking, which is the strongest cause of periodontitis, seems to be associated with higher tooth loss and with not receiving treatment,^2^ and this may have biased our estimate towards protective effects of periodontal therapy. Additionally, the adjustments made in the analyses may not fully account for the disease severity, which could influence both the likelihood of receiving periodontal treatment and experiencing tooth loss, potentially biasing our estimates towards no effect.^2^ On a slightly different note, a methodological challenge may have arisen from the fact that having a tooth extraction was a relatively infrequent event in the investigated population, and this results in an outcome variable with a rather skewed distribution, which is suboptimal for common statistical models.

When interpreting the findings it is important to bear in mind that periodontal therapy was defined as receiving any periodontal therapy within a calendar year. Clearly, this reflects the visiting intervals deemed appropriate for periodontitis patients by current dental community standards.^4,5^ However, this exposure definition may entail treatment patterns that some could consider suboptimal, e.g., periodontal therapy delivered at almost two-year intervals (visit in January, and then next in December in the following year). Furthermore, we cannot guarantee that the treatments provided align with contemporary standards.^4,5^ Accordingly, our treatment effect estimates may be biased towards null to an unknown extent. However, the implications of this underestimation are likely to be quite small because only a minority of people seem able or willing to receive periodontal therapy in the long term as recommended.^2,22^ Notwithstanding these limitations, the reported treatment effect estimates provide important insights about the real-world effectiveness of periodontal therapy on tooth loss in Denmark. Because there are no quality and long-term randomized clinical trials on the topic,^1^ analyzing rich and long-term routinely collected data has the potential to provide the meaningful evidence needed for clinical and regulatory decision-making. It is also necessary to evaluate and confirm the findings from smaller-scale clinical investigations, frequently conducted in hospital or specialist settings,^3^ which may not be well generalizable to the populations and settings where most of the periodontal care is delivered.

Unfortunately, the few earlier studies^23–25^ investigating the effectiveness of periodontal care on tooth loss using routinely collected data have had considerable limitations regarding the data, study design and analysis techniques required for credible causal inference using routinely collected healthcare data.^26,27^ A Taiwanese study^24^ compared 1.5-year cumulative incidence of tooth extractions between groups receiving either conventional or more comprehensive periodontal treatment course using health insurance data. After adjusting baseline demographic, socioeconomic, selected health conditions and clinic attributes, those who received comprehensive periodontal treatment course were less likely (OR: 0.83; 95% CI: 0.81; 0.85) to get tooth extraction over 1.5-year follow-up. Using health insurance data, a German group^23^ investigated the 4-year cumulative incidence of tooth extractions among a group which had received periodontal care periodontal care during previous 4 years and a group which had not received. The groups were matched based on age, sex and region. Those who had received treatment had slightly higher 4-year cumulative tooth extraction incidence (36.2% vs. 27.5%). Another German study^25^ compared tooth loss rates across three cohorts: periodontally treated patients from routine health insurance data, a population-based cohort (including both periodontally treated and untreated), and periodontally treated patients in a university hospital setting. The study examined unadjusted average yearly tooth loss rates and incidence rates of at least one tooth extraction across these cohorts. Results showed that the population-based cohort, regardless of whether received periodontal care or not, had lower tooth loss rates compared to both the register-based (insurance data) and university hospital cohorts. Comparing these conflicting findings to our results is difficult or even impossible, but it is worth mentioning that our study represents a significant methodological improvement in the analysis of routinely collected healthcare data in periodontology.

What then emerges from observational studies on the effectiveness of receiving periodontal care on tooth loss in more clinical settings? Based on eight relatively small clinical observational studies, it has been meta-analytically estimated that those who adhere to periodontal therapy would save approximately one tooth per eight years compared to those who adhere to periodontal therapy erratically.^3^ This estimate is considerably higher than that indicated by the findings of the present study. We estimated that a person receiving periodontal therapy in five calendar years in a row, would save an average of 0.06 (95% CI: 0.03; 0.11) teeth compared to a similar person who received no periodontal therapy during those five years. This effect is a quite small, considering that on average approximately 0.36 teeth (0.072 per year) were lost in a five-year period. Thus, in contrast to what has been expected in the literature,^7^ the rate of tooth loss seems also to be relatively modest not only in regularly treated but also in non-treated or erratically treated individuals. Notably, the estimated effect is based on a rather extreme contrast in the periodontal treatment pattern (no treatment in any of five years versus treatment in all five years), and thus we can assume that reducing periodontal treatment frequency, for instance, to every second year, would likely have an even smaller effect on tooth loss. Even though the decade before age 40 is associated with the highest periodontitis incidence,^28^ it should be borne in mind that the cohort studied here was relatively young at the outset, 40 years. While we found no impact of the severity of periodontitis on the relationship between periodontal therapy adherence and tooth extractions, it might be speculated that the level of severity of periodontitis has yet to reach a level where tooth extraction become a prominent treatment option. Certainly, it has been observed^29^ that the risk of tooth loss during periodontal maintenance is greater among the 60+ year-olds. In addition to differences in study design and definition of treatment adherence, a key difference between the present study and those included in the meta-analysis^3^ is that the present analysis adjusted for time-invariant and time-varying confounding, whereas the mentioned meta-analytical estimates were calculated based on unadjusted data. However, it is an untenable assumption that those who adhere to periodontal care are similar to those who do not. While it is well-known that those who receive periodontal therapy differ from those who do not in terms of their baseline characteristics,^2,30–32^ it is less well recognized that whether one opts for treatment at a certain time point is also influenced by the past periodontal treatment experience and tooth loss, leading to time-varying confounding with treatment-confounder feedback over time. In such situations, methods like g-estimation are required to properly estimate the treatment effect, and more traditional approaches are likely leading to biased treatment effect estimates, because the effects of previous treatments or outcomes on current treatment or future outcomes are not adequately adjusted for.^13,14,18–21^

In summary, this register-based analysis showed that receiving periodontal therapy has a slightly reducing effect on tooth extractions one to three years ahead. While acknowledging the constraints of our study, our findings offer meaningful evidence on the real-world effectiveness of periodontal therapy against tooth loss which seems to be considerably smaller than indicated by earlier clinical studies. Future research should explore the potential of utilizing routinely collected healthcare and patient-reported outcome measure data to estimate the impact of periodontal therapy on other patient-important outcomes, such as orofacial function, pain, appearance, and psychosocial well-being.^33^

## Acknowledgements

This work was partly supported by the Danish Rheumatism Association (Gigtforeningen) #R171-A5894.

## Author contributions

Eero Raittio: Conceptualization; Formal Analysis; Methodology; Investigation; Visualization; Writing – original draft. Vibeke Baelum: Conceptualization; Data curation; Investigation; Visualization, Project administration; Supervision; Writing – review & editing.

## Data availability statement

The data that support the findings of this study are available from Statistics Denmark. Restrictions apply to the availability of these data, which were used under license for this study.

## Declaration of generative AI and AI-assisted technologies in the writing process

During the preparation of this work the author(s) used GPT-4o in order to refine spelling and grammar. After using this tool/service, the author(s) reviewed and edited the content as needed and take(s) full responsibility for the content of the published article.

## References

1. Manresa C, Sanz-Miralles EC, Twigg J, Bravo M. Supportive periodontal therapy (SPT) for maintaining the dentition in adults treated for periodontitis. Cochrane Database Syst Rev 2018;1:CD009376. 10.1002/14651858.CD009376.pub2

2. Echeverria JJ, Echeverria A, Caffesse RG. Adherence to supportive periodontal treatment. Periodontol 2000 2019;79:200–209. 10.1111/prd.12256

3. Lee CT, Huang HY, Sun TC, Karimbux N. Impact of Patient Compliance on Tooth Loss during Supportive Periodontal Therapy: A Systematic Review and Meta-analysis. J Dent Res 2015;94:777–786. 10.1177/0022034515578910

4. Sanz M, Herrera D, Kebschull M, et al. Treatment of stage I-III periodontitis-The EFP S3 level clinical practice guideline. J Clin Periodontol 2020;47 Suppl 22:4–60. 10.1111/jcpe.13290

5. Herrera D, Sanz M, Kebschull M, et al. Treatment of stage IV periodontitis: The EFP S3 level clinical practice guideline. J Clin Periodontol 2022;49 Suppl 24:4–71. 10.1111/jcpe.13639

6. West N, Chapple I, Claydon N, et al. BSP implementation of European S3 - level evidence-based treatment guidelines for stage I-III periodontitis in UK clinical practice. J Dent 2021;106:103562. 10.1016/j.jdent.2020.103562

7. Trombelli L, Franceschetti G, Farina R. Effect of professional mechanical plaque removal performed on a long-term, routine basis in the secondary prevention of periodontitis: a systematic review. J Clin Periodontol 2015;42 Suppl 16:S221–236. 10.1111/jcpe.12339

8. Miyamoto T, Kumagai T, Jones JA, Van Dyke TE, Nunn ME. Compliance as a prognostic indicator: retrospective study of 505 patients treated and maintained for 15 years. J Periodontol 2006;77:223–232. 10.1902/jop.2006.040349

9. Salvi GE, Mischler DC, Schmidlin K, et al. Risk factors associated with the longevity of multi-rooted teeth. Long-term outcomes after active and supportive periodontal therapy. J Clin Periodontol 2014;41:701–707. 10.1111/jcpe.12266

10. Eickholz P, Kaltschmitt J, Berbig J, Reitmeir P, Pretzl B. Tooth loss after active periodontal therapy. 1: patient-related factors for risk, prognosis, and quality of outcome. J Clin Periodontol 2008;35:165-174. 10.1111/j.1600-051X.2007.01184.x

11. Listl S, Jurges H, Watt RG. Causal inference from observational data. Community Dent Oral Epidemiol 2016;44:409–415. 10.1111/cdoe.12231

12. Schuch HS, Nascimento GG, Demarco FF, Haag DG. Causal inference in dentistry: Time to move forward. Community Dent Oral Epidemiol 2023;51:62–66. 10.1111/cdoe.12802

13. Loh WW, Ren DN. A Tutorial on Causal Inference in Longitudinal Data With Time-Varying Confounding Using G-Estimation. Adv Meth Pract Psych 2023;6. 10.1177/25152459231174029

14. Loh WW, Ren DN. Estimating Time-Varying Treatment Effects in Longitudinal Studies. Psychol Methods 2023. 10.1037/met0000574

15. Vilstrup L, Christensen LB, Borge H, Kristensen SF. Oral health for users of private dental practice from 2000 to 2008 [in Danish]. Tandlaegebladet 2010;114:704–712.

16. Bolstad AI, Fevang BS, Lie SA. Increased risk of periodontitis in patients with rheumatoid arthritis: A nationwide register study in Norway. J Clin Periodontol 2023;50:1022–1032. 10.1111/jcpe.13826

17. Rosing K, Christensen LB, Damgaard C. Periodontal care attendance in Denmark in 2012-2016 - a nationwide register-based study. Acta Odontol Scand 2022;80:264–272. 10.1080/00016357.2021.1998611

18. Dukes O, Vansteelandt S. A Note on G-Estimation of Causal Risk Ratios. American Journal of Epidemiology 2018;187:1079–1084. 10.1093/aje/kwx347

19. Loh WW, Ren DN. The Unfulfilled Promise of Longitudinal Designs for Causal Inference. Collabra-Psychol 2023;9. 10.1525/collabra.89142

20. Tompsett D, Vansteelandt S, Dukes O, De Stavola BL. gesttools: General Purpose G-Estimation in R. Observational Studies 2022;8:1–28.

21. Vansteelandt S, Sjolander A. Revisiting g-estimation of the Effect of a Time-varying Exposure Subject to Time-varying Confounding. Epidemiologic Methods 2016;5:37–56. doi:10.1515/em-2015-0005

22. Knight ET, Murray Thomson W. A public health perspective on personalized periodontics. Periodontol 2000 2018;78:195–200. 10.1111/prd.12228

23. Raedel M, Priess HW, Bohm S, Noack B, Wagner Y, Walter MH. Tooth loss after periodontal treatment-Mining an insurance database. J Dent 2019;80:30–35. 10.1016/j.jdent.2018.11.001

24. Chan CL, You HJ, Lian HJ, Huang CH. Patients receiving comprehensive periodontal treatment have better clinical outcomes than patients receiving conventional periodontal treatment. J Formos Med Assoc 2016;115:152–162. 10.1016/j.jfma.2015.10.017

25. Kocher T, Holtfreter B, Priess HW, et al. Tooth loss in periodontally treated patients: A registry- and observation-based analysis. J Clin Periodontol 2022;49:749–757. 10.1111/jcpe.13668

26. National Institute for Health and Care Excellence. *NICE real-world evidence framework.* National Institute for Health and Care Excellence; 2022. p.

27. Hernan MA, Robins JM. *Causal Inference: What If*. Boca Raton: Chapman & Hall/CRC; 2020. p.

28. Kassebaum NJ, Bernabe E, Dahiya M, Bhandari B, Murray CJ, Marcenes W. Global burden of severe periodontitis in 1990-2010: a systematic review and meta-regression. J Dent Res 2014;93:1045–1053. 10.1177/0022034514552491

29. Chambrone L, Chambrone D, Lima LA, Chambrone LA. Predictors of tooth loss during long-term periodontal maintenance: a systematic review of observational studies. J Clin Periodontol 2010;37:675–684. 10.1111/j.1600-051X.2010.01587.x

30. Delatola C, Adonogianaki E, Ioannidou E. Non-surgical and supportive periodontal therapy: predictors of compliance. J Clin Periodontol 2014;41:791–796. 10.1111/jcpe.12271

31. Ramseier CA, Kobrehel S, Staub P, Sculean A, Lang NP, Salvi GE. Compliance of cigarette smokers with scheduled visits for supportive periodontal therapy. J Clin Periodontol 2014;41:473–480. 10.1111/jcpe.12242

32. Kocher T, Losler K, Pink C, Grabe HJ, Holtfreter B. Effect of Discontinuation of Supportive Periodontal Therapy on Periodontal Status-A Retrospective Study. J Clin Periodontol 2024. 10.1111/jcpe.14062

33. Mittal H, John MT, Sekulic S, Theis-Mahon N, Rener-Sitar K. Patient-Reported Outcome Measures for Adult Dental Patients: A Systematic Review. J Evid Based Dent Pract 2019;19:53–70. 10.1016/j.jebdp.2018.10.005

