## Supplementary material for "Assessing the impact of periodontal therapy on tooth loss: a register-based longitudinal study in Denmark"

**Supplementary information about variables used**

**Gender**: Men; Women. Information from: The Danish Civil Registration System (Pedersen 2011). Variable name in the register: KOEN.

**Origin**: Categorized into two groups: 1) Immigrants or descendants, and 2) persons of Danish origin. (Pedersen 2011). Variable name in the register: IE_TYPE

**Municipality of residence**: 99 municipalities. Information from: The Danish Civil Registration System (Pedersen 2011). Variable name in the register: KOM.

**Highest completed education**: categorized into eight groups:

•	Primary school

•	High school

•	Vocational

•	Short-cycle higher education or Qualifying exam

•	Medium-cycle higher education

•	Bachelor

•	Long cycle higher education or researcher education

•	Unknown

Information from the Danish Educational Register (Jensen et al. 2011). Categorized based on the variable in the register AUDD. Qualifying exam and Researcher education categories were combined with closest neighboring category due to their small size.

**Income percentile**: People were divided into 100 groups (percentiles) for each year from 1990 to 2021 based on their total annual personal income. Personal income in total is equal to the sum of business income, transfer income, property income (excluding calculated rental value of own home) and other non-classifiable income that can be attributed directly to the individual. The amount is before tax deduction, labor market contribution and special pension contribution, and interest expenses are not deducted. Information from the Danish Income Statistics Register (Baadsgaard et al. 2011). Variable name in the register: PERINDKIALT_13.

**Number of dental restorations per calendar year**

From the National Health Insurance Service Register (Sahl Andersen et al. 2011), we used the dental treatment codes that have been used over time since 1990 in the National Health Insurance scheme covering the subsidized dental care for all adult permanent residents in Denmark. A total number of treatment codes representing amalgam or composite restorations (1501, 1502, 1503, 1504, 1505, 1506, 1507, 1509, 1551, 1552, 1553, 1554, 1555, 1556, 1557, 1558, 1559) were summed in each calendar year.

**Incident diabetes mellitus type 1 or 2:** Data on incident diabetes between 1997 and 2021 comes from the Registry for Selected Chronic Diseases and Severe Mental Disorders (Danish Health Data Authority 2024). It combines information from the Danish National Prescription Registry (Pottegard et al. 2017), including all prescriptions in Denmark, and the Danish National Patient Registry (Schmidt et al. 2015) which contains information on all visits, procedures, and admissions to all Danish somatic hospitals, emergency departments, and hospital-associated outpatient clinics.

Individuals were classified as having incident diabetes mellitus (type 1 or 2) in a calendar year when one of the following criteria were met.

Diabetes type 1:

- Individuals registered with at least two purchases of insulin or insulin analogs (A10A, except combination medicines including GLP1-analogues and insulins, A10AE54 or A10AE56) in the Danish National Prescription Registry.
- Individuals registered with a relevant primary or secondary diagnosis (E10, diabetes type I, or its sub-codes under ICD-10) in the Danish National Patient Registry.

Diabetes type 2:

- Individuals registered with at least two purchases of medication aimed at lowering blood glucose (A10B, except A10BJ, A10BK01, A10BK03) or combination medicines including GLP1-analogues and insulins (A10AE54 or A10AE56) in the Danish National Prescription Registry.
- Individuals registered with a relevant primary or secondary diagnosis (E11, diabetes type 2, or its sub-codes under ICD-10) in the Danish National Patient Registry.

Excluded were:

- Women who have been exclusively treated with metformin (ATC code A10BA02) and there were signs that they could have polycystic ovary syndrome (prescription for G03GB02, G03HB or diagnosis code E282).
- Women who have a code for gestational diabetes (ICD-10 code O24.4) and who have only registered purchase of antidiabetics (A10) within 280 days before first contact or 280 days after last contact with gestational diabetes according to the Danish National Patient Registry.

**Number of teeth, number of filled teeth, and number of decayed teeth**

Since 2000, private dental practitioners have had to report the number of teeth, number of filled teeth, and number of decayed teeth for their patients turning 25, 40 or 65 years during the ongoing calendar year to the National Board of Health (Vilstrup et al. 2010) where these data have become integrated with the National Health Insurance Service Register. We extracted data on the number of teeth, number of filled teeth, and number of decayed teeth.

Data was cleaned as follows. If the recorded number of teeth was 0 (unlikely in 40-year-old Danes in 2001) or exceeded 32, data was considered as missing. Similarly, if recorded number of filled or number of filled decayed teeth exceeded 32, data was considered as missing. For all three variables values in the range from 29 to 32 were replaced with 28.

**G-estimation in general**

Using the Figure 3 as an example, the simplified outline of the g-estimation process (Vansteelandt et al. 2016; Dukes et al. 2018; Tompsett et al. 2022; Loh et al. 2023c; 2023b; 2023a) goes as follows: Estimation proceeds by focusing in turn on each outcome time (O2 and O3) and sequentially estimating the treatment effects from previous times (E1 and E2) on that outcome. Importantly, the latest outcome, O3, can be considered as the result of accumulation of the treatment effects from all previous time points (E1 and E2). ”Peeling off” the effect of E2 on O3 from the variation in O3 allows unbiased estimation of the effect of E1 on O3 by a simple outcome (O3) regression model including E1 and all pre-exposure variables (C, W1) while holding E2 fixed at a reference level (not treated) for everyone (Vansteelandt et al. 2016; Dukes et al. 2018; Loh et al. 2023b; 2023a). To make these outcome models more robust, they are supplemented with balancing propensity scores for confounders, previous exposures, and previous outcome status for each exposure time (E1 and E2). For instance, the outcome model estimating the effect of E1 on O3 includes propensity scores to balance C and W1 by E1 status. Therefore, g-estimation can be considered a doubly robust strategy which is not so dependent on a correctly specified single propensity score or outcome model (Vansteelandt et al. 2016; Dukes et al. 2018; Tompsett et al. 2022; Loh et al. 2023c; 2023b; 2023a). G-estimation models can also be supplemented with weights from a censoring model to control for potentially differing drop-out probabilities depending on previous exposure, outcome, and confounder values (Vansteelandt et al. 2016; Dukes et al. 2018; Tompsett et al. 2022; Loh et al. 2023c; 2023b; 2023a).

Even though being a sophisticated approach, three common causal inference assumptions need to hold in order to correctly identify the causal effect with g-estimation; conditional exchangeability, that is no unmeasured confounding; positivity, that is above zero probability of receiving exposure; and counterfactual consistency, which dictates that an “individual’s counterfactual outcome under a specific set of exposures is equal to their outcome had it been their observed exposure history.”(Tompsett et al. 2022)

Rather than reporting all three effect estimates from the DAG example (E1 on O2, E1 on O3, and E2 on O3) - which with 10 time points could amount to 55 effects (1+2+…+10=55) - one can estimate the effect of exposure on the outcome up to a specified number of time points between exposure and outcome (Vansteelandt et al. 2016; Tompsett et al. 2022). In the example, the effect estimates for E1 on O2 and E2 on O3 represent the effect of E on O one time point ahead, whereas the effect estimate E1 on O3 represents the effect of E on O two time points ahead. This offers several advantages, including improved computational efficiency by focusing on the most relevant and accurately estimated effects. It also allows for the exploration of how the effect of E on O may change over time, potentially increasing or decreasing as the interval between them widens. Furthermore, when O is a continuous outcome, the sum of these time-varying effects can be interpreted as the average treatment effect on tooth extractions of receiving periodontal therapy at all time points versus not receiving it at any time point during follow-up (Vansteelandt et al. 2016; Dukes et al. 2018; Tompsett et al. 2022).

**Lag effects from previous exposure, outcome and confounders**

The rationale behind the decision to allow a 1-to-5-year lag, rather than using all historical data, was to maintain computational efficiency without changing the primary estimate considerably. Principally, using the notation in the DAG (Figure 3), allowing a 1-to-5-year lag effect means that e.g., when investigating the effect of periodontal therapy in the sixth year, E6, on the number of extractions in the seventh year, O7, both the outcome model and the propensity score models include C, W6, V0-V5, E0-E5 and O0-O5. The same logic applies for all propensity score and outcome models in all other years. It is also worth noting that our approach allowed account of 1-to-5-year lag effects for E, O and V already from the beginning of the follow-up (2002, T=1) and onwards.

**Performing G-estimation with** **gesttools package**

First, data was formatted according to guidance given for using gesttools package (Tompsett et al. 2022). Analyses were performed with gestMultiple function and bootsrapped with gestboost functions using following specifications. Outcome, propensity score, and censoring models included variables listed in Table S1. Outcome models also naturally included the exposure, receiving periodontal therapy in a calendar year. Time-varying effect (“type=3”) on tooth extractions five years ahead (“cutoff=5”) were estimated.

Effect medication by disease severity status was estimated by including interaction of severe periodontitis at baseline and receiving periodontal therapy in the outcome models. Time-varying effects allowing for effect modification (“type=4”) on tooth extractions five years ahead (“cutoff=5”) were estimated.

By default, the gesttools package works with binary and continuous outcomes (Tompsett et al. 2022). However, because count and binary outcomes can be estimated with the same procedure using gamma regression for outcome models (Dukes et al. 2018), we made minor modifications to the gesttools functions so that they can be used for count outcomes. R-scripts for these modified functions are available from GitHub and supplemented with validation analyses (<https://github.com/raittioe/gestCount>).

**Sensitivity analyses**

In the sensitivity analyses assuming the alternative directed acyclic graph (Figure 4), when estimating the effect from E6 to O7, the adjustment sets in propensity score and outcome models would change from C, W6, V0-V5, E0-E5 and O0-O5 to C, W6, V1-V6, E1-E5 and O1-O6, and similarly for all other combinations of exposure and outcome times.

**References**

Baadsgaard M, Quitzau J. 2011. Danish registers on personal income and transfer payments. Scand J Public Health. 39(7_suppl):103-105.

Danish Health Data Authority. 2024. Registry for selected chronic diseases and severe mental disorders [in danish].

Dukes O, Vansteelandt S. 2018. A note on g-estimation of causal risk ratios. Am J Epidemiol. 187(5):1079-1084.

Jensen VM, Rasmussen AW. 2011. Danish education registers. Scand J Public Health. 39(7_suppl):91-94.

Loh WW, Ren DN. 2023a. Estimating time-varying treatment effects in longitudinal studies. Psychol Methods.

Loh WW, Ren DN. 2023b. A tutorial on causal inference in longitudinal data with time-varying confounding using g-estimation. Adv Meth Pract Psych. 6(3).

Loh WW, Ren DN. 2023c. The unfulfilled promise of longitudinal designs for causal inference. Collabra-Psychol. 9(1).

Pedersen CB. 2011. The danish civil registration system. Scand J Public Health. 39(7_suppl):22-25.

Pottegard A, Schmidt SAJ, Wallach-Kildemoes H, Sorensen HT, Hallas J, Schmidt M. 2017. Data resource profile: The danish national prescription registry. 46(3):798-798f.

Sahl Andersen J, De Fine Olivarius N, Krasnik A. 2011. The danish national health service register. Scand J Public Health. 39(7_suppl):34-37.

Schmidt M, Schmidt SA, Sandegaard JL, Ehrenstein V, Pedersen L, Sorensen HT. 2015. The danish national patient registry: A review of content, data quality, and research potential. 7:449-490.

Tompsett D, Vansteelandt S, Dukes O, De Stavola BL. 2022. Gesttools: General purpose g-estimation in r. 8:1 - 28.

Vansteelandt S, Sjolander A. 2016. Revisiting g-estimation of the effect of a time-varying exposure subject to time-varying confounding. 5(1):37-56.

Vilstrup L, Christensen LB, Borge H, Kristensen SF. 2010. Oral health for users of private dental practice from 2000 to 2008 [in danish]. 114(9):704-712.

**Supplementary tables and figures**

Table S1. Odds ratios (ORs) for receiving periodontal therapy within a calendar year from a logistic regression model.

OR	2.5%	97.5%	z val.

Intercept	0.07	0.04	0.11	-9.58

Severe periodontitis at baseline (ref. No)

Yes	1.10	1.02	1.18	2.47

Gender (ref. Women)

Men	0.82	0.75	0.88	-5.16

Danish origin (ref. No)

Yes	1.31	1.14	1.50	3.86

Region (ref. North Jutland)

Central Jutland	0.85	0.73	0.98	-2.19

Southern Denmark	0.87	0.75	1.01	-1.84

Zealand	0.84	0.73	0.97	-2.38

Greater Copenhagen	0.87	0.75	1.02	-1.69

Number of teeth at baseline	1.01	1.00	1.02	1.53

Number of filled teeth at baseline, splines

1^st^ spline	0.86	0.72	1.02	-1.77

2^nd^ spline	0.89	0.63	1.28	-0.62

3^rd^ spline	1.24	0.94	1.64	1.53

Number of decayed teeth at baseline	0.98	0.97	0.99	-3.85

Highest completed education (ref. Unknown)

Primary school	1.44	1.04	2.00	2.18

High school	1.60	1.11	2.29	2.53

Vocational	1.65	1.19	2.29	3.01

Short-cycle higher education or Qualifying exam	1.80	1.26	2.57	3.22

Medium-cycle higher education	1.75	1.25	2.46	3.22

Bachelor	1.25	0.81	1.94	1.00

Long cycle higher education or researcher education	1.83	1.27	2.66	3.21

Income percentile, splines

1^st^ spline	1.57	1.35	1.83	5.79

2^nd^ spline	1.64	1.11	2.42	2.48

3^rd^ spline	1.40	1.21	1.62	4.58

Diabetes, type 1 or type 2 (ref. No)

Yes	0.97	0.80	1.16	-0.36

Periodontal therapy in previous years

Lag1	5.52	5.07	6.01	39.44

Lag2	2.55	2.32	2.79	19.90

Lag3	1.90	1.73	2.09	13.14

Lag4	1.54	1.40	1.70	8.71

Lag5	1.77	1.61	1.94	12.05

Number of restorations in previous years

Lag1	1.03	1.01	1.06	2.48

Lag2	1.03	1.00	1.06	2.10

Lag3	1.01	0.99	1.04	1.04

Lag4	0.97	0.95	1.00	-2.00

Lag5	0.97	0.95	0.99	-2.42

Number of extractions in previous years

Lag1	1.03	0.97	1.09	0.99

Lag2	0.95	0.89	1.01	-1.75

Lag3	0.92	0.86	0.98	-2.50

Lag4	0.95	0.89	1.02	-1.38

Lag5	0.93	0.86	1.00	-1.98

Year (ref. 2002)

2003	0.73	0.58	0.91	-2.83

2004	0.83	0.66	1.04	-1.66

2005	0.82	0.66	1.03	-1.70

2006	0.60	0.48	0.75	-4.50

2007	0.95	0.76	1.19	-0.42

2008	0.98	0.79	1.23	-0.15

2009	0.91	0.73	1.14	-0.80

2010	0.82	0.65	1.02	-1.78

2011	0.99	0.79	1.25	-0.06

2012	0.85	0.68	1.06	-1.43

2013	0.50	0.40	0.62	-6.24

2014	0.63	0.50	0.79	-4.04

2015	0.92	0.73	1.15	-0.76

2016	0.88	0.70	1.10	-1.14

2017	0.91	0.73	1.15	-0.77

2018	0.88	0.70	1.11	-1.09

2019	0.85	0.68	1.07	-1.38

2020	0.61	0.49	0.77	-4.24

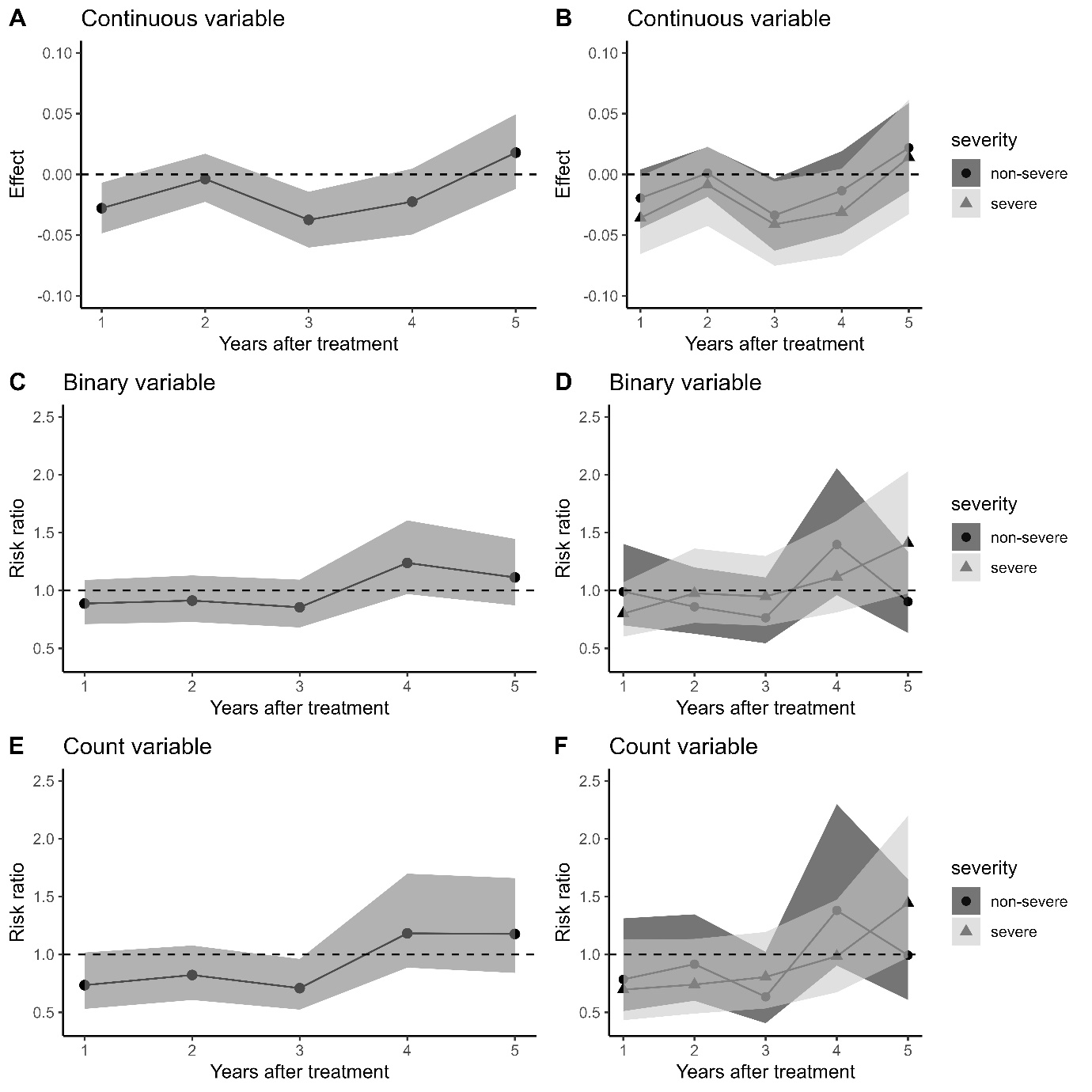

Figure S1. The effect of receiving periodontal treatment within a calendar year on the number of extracted teeth (A, B, E, F) or probability of at least one tooth extraction (C, D) one to five years later. The right panel shows the effect of receiving periodontal treatment within a calendar year in those with sever or non-severe condition at the baseline. **Adjustment set selected based on the alternative DAG (Figure 4).**
